# Implicit and explicit motor learning interventions for gait in people after stroke: a process evaluation of a randomized controlled trial

**DOI:** 10.1101/2020.01.17.20017897

**Authors:** Li-Juan Jie, Melanie Kleynen, Kenneth Meijer, Anna Beurskens, Susy Braun

## Abstract

**Background:** Gait training within stroke rehabilitation can be applied using implicit or explicit motor learning approaches. Explicit learning is a more conscious approach to learning, in which many detailed instructions about the movement are provided to the learner. Implicit learning strives to take place in a more automatic manner, without much knowledge of the underlying facts and rules of the movement.

**Objective:** To evaluate whether the implicit and explicit motor learning walking interventions for people after stroke delivered in a randomized controlled trial were performed as intended (fidelity) and to report the therapist and participant experiences with regard to feasibility.

**Methods:** Fidelity was assessed by evaluating the dose delivered (number of therapy sessions) and content of instructions (explicit rules) that were collected through the therapist logs and audio recordings of the training sessions. The therapist and participant experiences were assessed by means of self-developed questionnaires.

**Results:** 79 people were included of which seven people (9%) dropped out. The remaining participants all received the required minimum of seven sessions. Overall therapists adhered to the intervention guideline. On average 5.2 and 0.4 explicit rules were used within the explicit and implicit group respectively. Therapists and participants were generally positive about the feasibility but frequent comments were made by the therapists regarding “therapy time restrictions” and “tendency of the participants to develop explicit rules”.

**Conclusion:** Delivery of the implicit and explicit motor learning walking interventions were successful in terms of fidelity. Therapists and participants were generally positive about the feasibility of the intervention.

## Introduction

Within stroke rehabilitation gait is one of the most practiced activities [1]. One way to apply over ground gait training is to use an explicit or implicit motor learning approach. Explicit learning is typically referred to as a more conscious approach to learning [2], in which many detailed instructions about the movement itself are provided to the learner. Whereas implicit learning strives to take place in a more automatic manner, without much knowledge of the underlying facts and rules of the movement [2]. To apply implicit motor learning different learning strategies can be used, for example errorless learning, dual task learning or analogy learning. In analogy learning one single analogical rule that integrates all (explicit) knowledge to perform the to-be-learned motor skill is used. For example, instead of providing many detailed instructions (explicit rules) to increase step length, therapists could also consider to use an (personalised) analogy such as “follow the footprints in the sand” [3]. Looking at current clinical practice physiotherapists tend to use primarily a large amount of verbal instructions (explicit learning) or use a mix of implicit and explicit motor learning approaches [4,5]. However, within sport various advantages and positive results of implicit motor learning over explicit motor learning have been found, such as: better performance while multitasking or in pressurised environments and stable performance over a longer time period [6]. These positive results have the potential to transfer to the therapeutic environment. Furthermore, as implicit motor learning strives to minimise cognitive load in working memory [6], it may be a particularly interesting learning approach for those who suffer from cognitive impairments e.g., which is commonly seen in people after stroke.

To date the majority of studies on the effectiveness of implicit motor learning within the stroke population have been performed within laboratory settings and with non-functional computer tasks [7]. Only few pilot and feasibility studies have been performed within clinical practice using real life tasks. These studies demonstrate the feasibility and potential effects of implicit motor learning within neurological rehabilitation [3,8]. Given the limited evidence, a large randomized controlled trial [9] (preprint version) including a total of 79 participants was conducted, in which implicit and explicit walking interventions for people after stroke were compared. We hypothesized that implicit motor learning would result in greater improvements of walking speed post intervention [9]. It was found that participants in both intervention groups improved with little to no differences between the interventions. To our knowledge this was the first study that took place within a clinical setting and with personalised interventions. No rigid protocol was implemented, however a framework was provided for the implicit and explicit interventions. The process evaluation took place parallel to the randomized controlled trial and covered the following research questions:

- To what extent did the therapists deliver the interventions as intended (fidelity)?
- How did the therapist’s experience delivering the interventions with regard to the feasibility?
- How did the patients’ experience the intervention with regard to the feasibility?

## Method

In this prospective process evaluation both quantitative and qualitative methods were used. The protocol of the randomized controlled trial [10] was approved by the local Ethics committee METC-Z in Heerlen, the Netherlands (Ethics nr: 17-T-06) and registered in the Netherlands Trial Register (Trial NL6133 (NTR6272)).

### Participants, and therapists

Both the participants of the study and therapists who provided the interventions were included in the process evaluation. The participants were recruited through local private practices, rehabilitation institutes, and a local health-related newspaper. All the participants evaluated their experience about the received intervention through a participant questionnaire developed to be specific to the intervention.

The therapists were recruited through the (professional) network of the researchers. Only experienced therapists (> 10 years) within neurological rehabilitation and/or experts within field the of the motor learning were included in the study. All therapists provided both the implicit and explicit interventions.

### Interventions, training of therapists and assessors

Within this RCT an implicit and explicit motor learning walking intervention for people after stroke was evaluated. Based on scientific evidence [2,3,8,11] and expert experiences (practice based evidence) an intervention guideline was developed that outlined precisely how the treatments (implicit and explicit conditions) were supposed to be delivered. The guideline for both interventions was comparable in terms of duration (three sessions per week over a 3-week period and 30 minutes per session). A minimum total of seven sessions was considered as being consistent with the protocol [10]. Outside guided therapy and after the 3-week intervention period participants were asked to use the practiced instructions in daily life. First the treating therapist identified the individual walking problem (similar for both groups), after which tailored instructions were provided in either an implicit or explicit manner according to the intervention guideline. Figure 1 provides an overview of the main characteristics of the interventions. Below a summary of the provided implicit and explicit motor learning walking interventions is provided, more detailed descriptions have been reported elsewhere [9,10].

**Figure 1.**
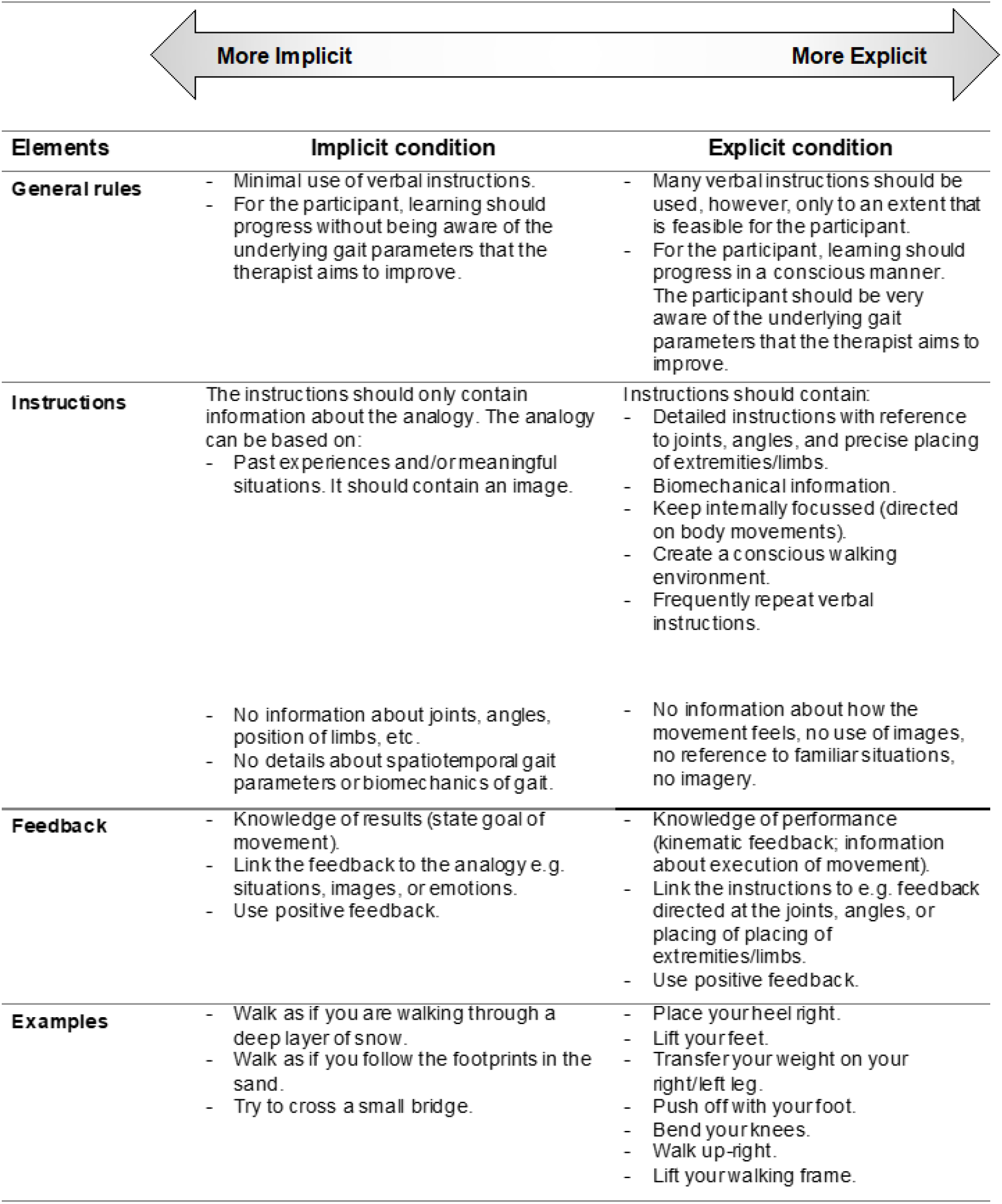
Overview with the main characteristics of the interventions. Adapted from Jie et al[10] under the terms of Creative Commons Attribution 4.0 license.

#### The implicit intervention

Within the implicit intervention we aimed to influence the walking pattern by facilitating the learning process with no or minimal increase of verbal knowledge [2]. Analogy learning was chosen to guide the implicit intervention. In analogy learning, complex information e.g. step-by-step rules (explicit knowledge) about the motor skill, is replaced by one simple biomechanical metaphor or analogy [12]. The therapists and patients had a booklet with example analogies as a source of inspiration. Based on the walking problem and participant preferences an analogy was created, practiced and adjusted if necessary, e.g. if the analogy did not lead to the desired adaptations of the movement performance. Additionally, therapists were allowed to use all elements that are reported in the implicit column displayed in figure 1.

#### The explicit intervention

Within the explicit intervention the opposite learning environment was created as much as possible, by organizing a conscious learning environment in which many verbal explicit instructions (explicit knowledge) were used. The therapists and patients had a card that displayed the different phases of the gait cycle that could to discus and provide detailed instructions and feedback on e.g., about the joints, angles or placements of the limbs. Although the explicit intervention strived to use as many instructions as appropriate, the number of instructions were used up to an amount that was considered to be possible (feasible) for the participants to process/handle. The latter was based upon as the judgement of the treating therapists. Additionally, therapists were allowed to use all elements that are reported in the explicit column displayed in figure 1.

To standardize both interventions, the therapists were trained during five sessions prior to the study. During these sessions the intervention guideline was discussed, and example cases were used to practice the application of the interventions. During the study three evaluation sessions took place to evaluate experiences or discuss difficulties and to ensure all therapists were using the intervention guideline in a similar manner.

### Data collection

Demographic information of the participants such as age, gender and time post stroke was gathered at baseline. To describe the characteristics of the population the following measures were assessed before the start of the interventions. First, global cognitive function was assessed using the Montreal Cognitive Assessment [13]. Second the Berg Balance Scale, was used to measure static and dynamic balance abilities [14]. The Rivermead Mobility Index, a short 10-item questionnaire was administered to assess mobility in gait, balance and transfers after stroke [15]. Furthermore, the Fugl-Meyer assessment for the lower limb was assessed to measure the participants ability to make movements outside the synergetic patterns [16]. Finally, the Movement Specific Reinvestment Scale (MSRS) was administered [17,18]. The MSRS is a short questionnaire with two factors, one related to conscious control of movements (conscious motor processing) and one related to self-consciousness about moving (movement self-consciousness) was assessed. Each factor of the MSRS comprises five statements, such as ‘I try to think about my movements when I carry them out’ (conscious motor processing) and ‘I am concerned about what people think about me when I am moving’ (movement self-consciousness). Table 1 presents an overview of the different measures, timing of the measurement assessments, number of participants who were received and completed the measurements that were used within the process evaluation.

**Table 1.**
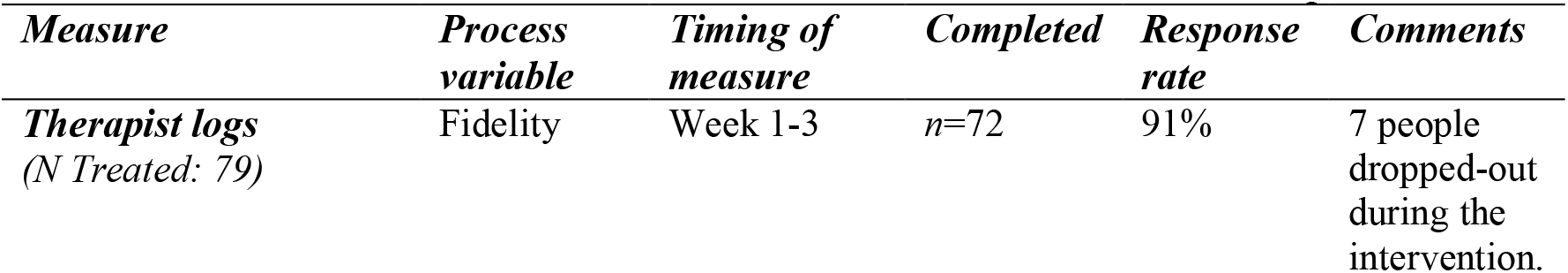
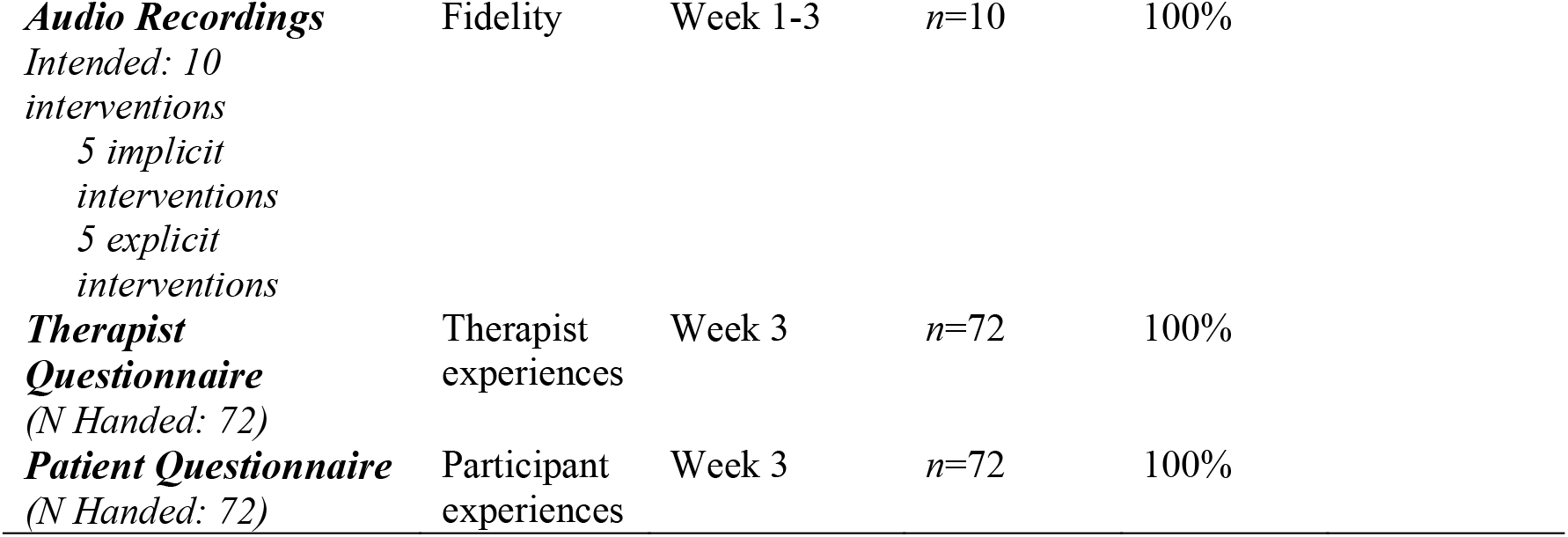
Overview of the different measures used to answer the research questions.

#### Fidelity

Within this process evaluation fidelity was assessed with regard to the dose (number of sessions) and content of instructions (adherence to the intervention guideline and the number of explicit rules provided). Data from the therapist logs, and audio files were used for this purpose. Therapist logs were completed immediately after each session and were used to evaluate therapists adherence with the intervention guideline [10]. In the logs the number of sessions delivered, the provided instructions and any deviations from the protocol were reported. In addition to the self-reported data (subjective) 10 gait training sessions (5 implicit and 5 explicit interventions) were selected at random during intervention period to be audio-recorded. However, therapists were aware that the recordings took place.

#### Therapist experiences with regard to feasibility

The therapist questionnaire included four questions, one question each to explore their thoughts on the intervention period (3-weeks in total), duration of the training sessions (30 minutes per session), the amount of sessions (9 in total) and future use of the learned strategy by the participant. The second part of the therapist questionnaire included four questions related to independent practice of the participants, suitability of the interventions, the tendency to develop explicit rules, and any observed changes or improvements in gait. The first four questions were recorded using binary outcomes ‘yes/no’, whereas the last four questions were scored on a 11-point Likert scale. There was space for free comments to elaborate on each response (see Table 4 for questions).

#### Patient experiences with regard to feasibility

The patient questionnaire was administered at the end of the nine training sessions. Participants evaluated the following three questions: 1) the ease-of-use of the provided instructions 2) their gained confidence in walking and 3) their overall experience with regard to the received gait training sessions. Each participant scored their satisfaction about the intervention on an 11-point Likert scale (see Table 5 for the questions).

### Data analysis

Data from the therapist logs (dose delivered, provided instructions), audio recordings (provided instructions), therapist- (feasibility) and participant- (feasibility) questionnaires were extracted by a research assistant and summarised in an excel spreadsheet. The quantitative outcomes of the questionnaires were presented using descriptive statistics (median, IQR). Frequently reported responses were described and quotes were used as clarifying examples.

The actual dose delivered was determined by counting the number of completed sessions within the therapist logs. The content (provided instructions) of the therapist logs and audio recordings were examined for the presence of explicit rules in both groups and the use of analogies within the explicit intervention by two independent raters [10]. To determine contrast between groups the number of explicit rules per participant was counted. To explore whether cognition influenced the quantity of explicit rules, the number of explicit rules were counted in people with lower cognitive abilities (MoCA ≤ 21) and vice versa. If explicit rules were used within the implicit intervention or when analogies were used within the explicit intervention this was reported as deviations from the protocol if they occurred in two or more sessions per participant [10]. To provide more insight in the content of the given instructions, an overview of examples within the implicit (analogy instructions) and explicit (explicit rules) interventions were presented.

## Results

In total 79 people after stroke were included in the main analyses of the study. During the intervention period seven participants (9%) dropped out (implicit group n=3; explicit group n=4). Three people wanted to improve overall fitness but did not have specific goals related to gait and two participants had other complaints not related to gait and therefore discontinued with the intervention. Furthermore, one participant stopped due to pregnancy and another person due to a change in medical diagnoses. These participants and their therapists did not evaluate the intervention (therapist and patient logs). Therefore, a total of 72 cases were included and evaluated in the process evaluation (Table 2). Based upon the three evaluation sessions with the treating therapists no deviation in the application of the intervention guideline were reported.

**Table 2.** Baseline characteristics of the participants (n=72).

|  | Implicit<br>(n = 35) | Explicit<br>(n = 37) |
| --- | --- | --- |
| **General Characteristics** |  |  |
| Age (yr), mean (SD) | 64.6 (9.4) | 67.8 (11.6) |
| Sex, n males (%) | 24 (63%) | 25 (61%) |
| Height (cm), mean (SD) | 171.7 (8.0) | 172.0 (8.8) |
| Educational level, n (%) |  |  |
| Elementary education | 0 (0%) | 1 (2%) |
| Secondary education | 20 (53%) | 18 (44%) |
| Vocational training | 10 (26%) | 11 (27%) |
| University | 8 (21%) | 11 (27%) |
| **Stroke Characteristics** |  |  |
| Time post stroke (months), mean (SD) | 72.8 (59.3) | 67.5 (69.1) |
| Side of stroke, n right (%) | 19 (50%) | 16 (19%) |
| Independent walking, n (%) |  |  |
| Walk unaided | 9 (24%) | 9 (22%) |
| Walk with stick | 29 (67%) | 32 (78%) |
| **Motor Characteristics** |  |  |
| BBS (0-56), mean (SD) | 45.5 (11.6) | 43.5 (8.9) |
| RMI (0-15), mean (SD) | 11.6 (2.5) | 11.3 (2.7) |
| FMA (0-34), mean (SD) | 23.5 (8.0) | 22.2 (8.1)* |
| **Cognitive Characteristics** |  |  |
| MoCA (0-30), mean (SD) | 24.7 (4.2) | 23.2 (6.2) |
| **Conscious motor control preference** |  |  |
| MSRS (0-10), mean (SD) | 4.9 (2.5) | 5.1 (2.6) |

### Fidelity

In terms of dose delivered the therapist’s logs revealed that in total 627 therapy sessions were delivered (303 implicit sessions; 324 explicit sessions). All 72 participants (100%) attended seven or more sessions according to protocol, of which 55 participants (76%) attended all nine, 13 participants (18%) completed eight and four participants (6%) completed seven training sessions.

In 4 (1%) of the 324 sessions within the explicit intervention, an analogy (implicit instruction) was used. In 14 (5%) of the 303 sessions within the implicit intervention, explicit rules were used. Two protocol deviations were observed within the implicit intervention i.e. explicit rules that were used in ≥ 2 sessions per participant. No protocol deviations were observed within the explicit condition. However, in one occasion, no explicit rules were used while being allocated to the explicit intervention. For this case the therapist log revealed that more general instructions were provided such as accelerate, decelerate or walk and look right, but these were not counted as explicit rules as they did not comply to the definition of explicit rules as defined within research protocol [10].

Therapist logs revealed that on average *M*=5.2 (*Range =* 0 - 10) explicit rules were used within explicit group and *M*=0.4 (*Range =* 0 - 3) explicit rules the implicit group. Within the explicit group, on average less explicit rules were used (*M* = 4.1, *R* = 2 – 8, *n* = 10) when participants were more cognitively impaired (MoCA < 21), compared to those who were not (*M* = 5.6, *R* = 0 – 10, *n* = 27). Table 3 provides an overview of the used instructions within the implicit and explicit group.

**Table 3.**
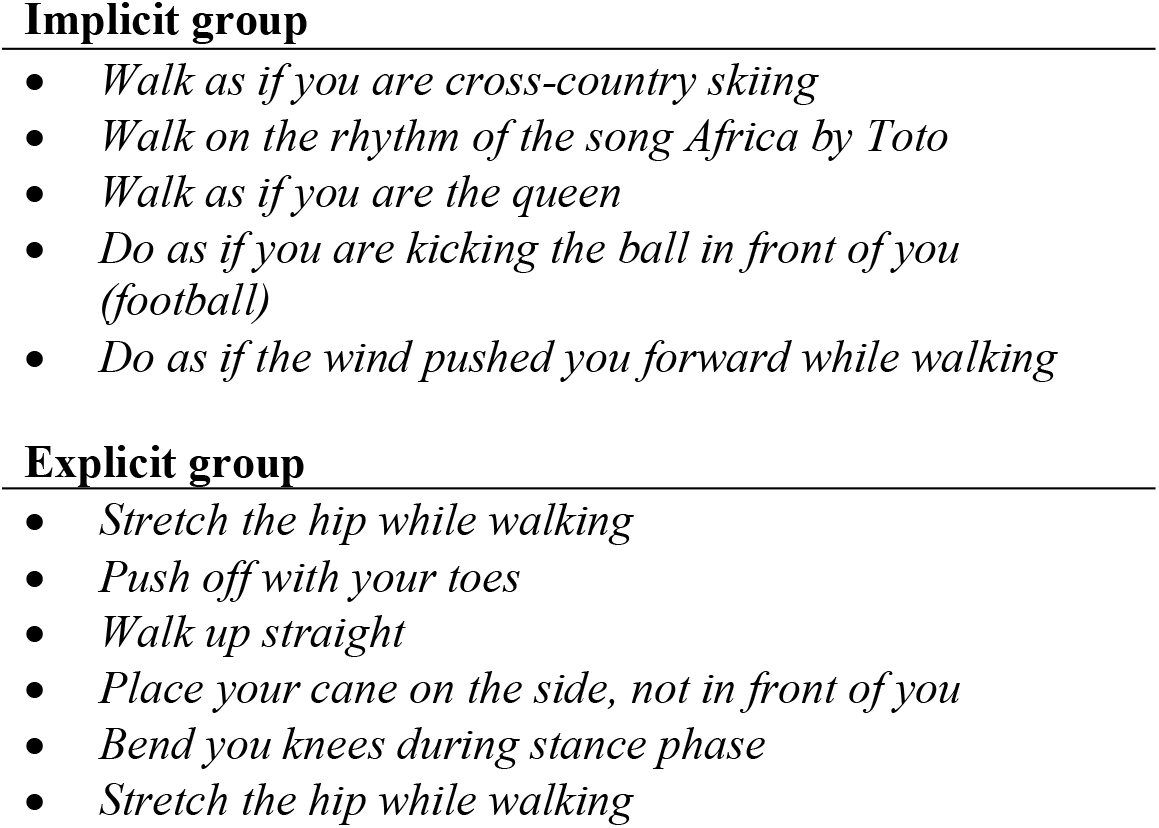
Examples of the provided instructions by the therapists.

### Therapist experiences with regard to feasibility

In table 4 subjective responses of the therapists related to the feasibility and observed changes of the intervention are presented. In the therapist questionnaire no apparent differences between the groups were visible (Table 4). Comments were frequently made with regard to “time restrictions”, “tendency to develop explicit rules” and “observed changes”.

**Table 4.**
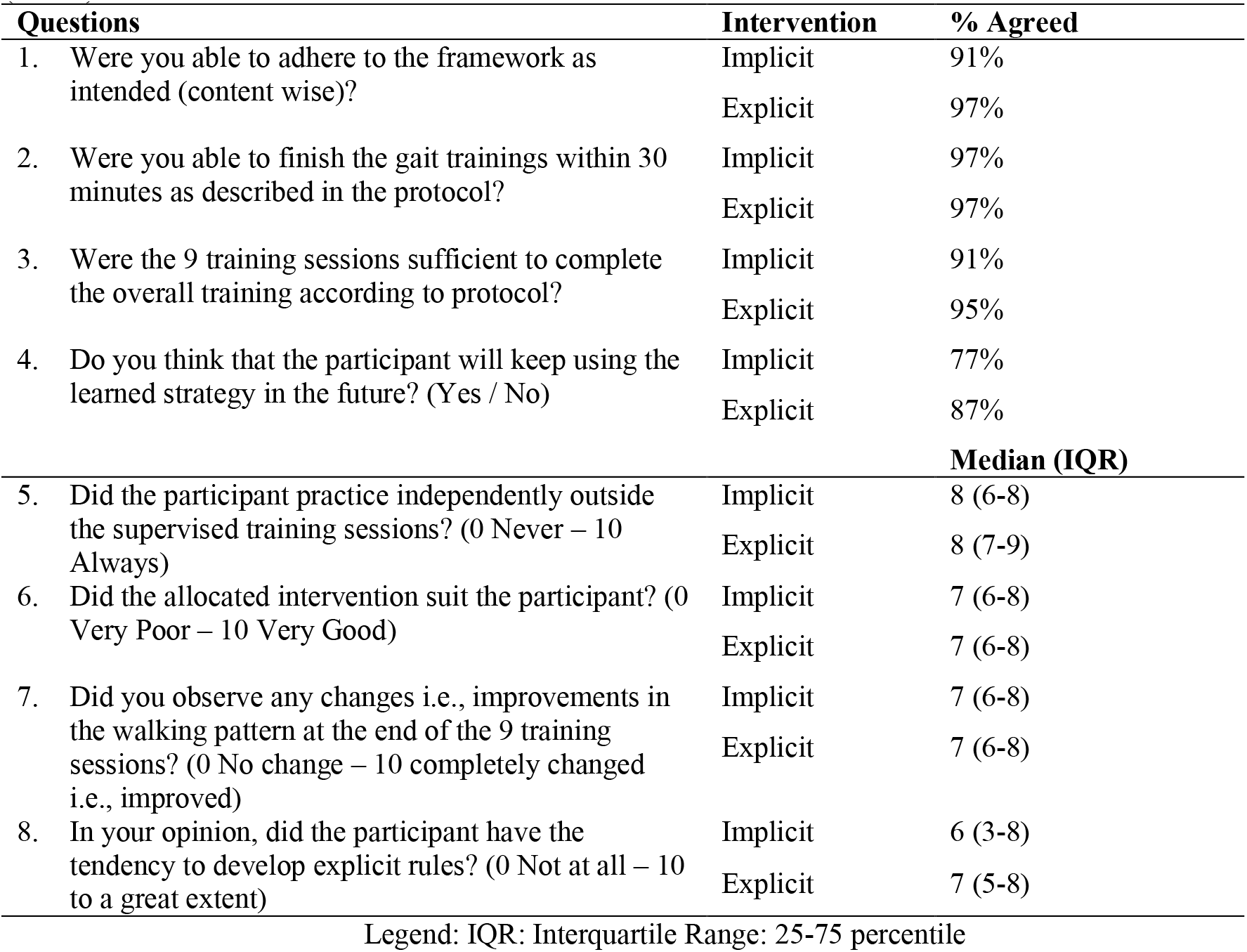
Responses of the therapist questions about their experiences and observed changes (*n*=72).

#### Time restrictions

Despite good adherence to the protocol (questions 1 to 3), therapists in both groups mentioned more time was preferred (> nine sessions and > 30 minutes per session) when participants were aphasic. Furthermore, when therapists provided the implicit intervention, they mentioned that sometimes more than one therapy session was preferred to establish a personalized metaphor. For example, therapists mentioned *“I needed the first four sessions find the right approach. With music, and rhythm music I experienced good results. Three extra sessions would have been effective for him*” (implicit intervention, therapist 3), or “*The first session, 30 minutes were tight. Later on, 30 minutes seemed sufficient*” (implicit intervention, therapist 3).

#### Tendency to develop explicit rules

Related to the development of explicit rules (question 8), therapists mentioned that often the participants wanted to be in control (think about) of their own walking performance. Therapists described “*as a person she wants to control*” (explicit intervention, therapist 4) or “*He thinks a lot about ‘why’ and ‘what” is going wrong during walking*’ (implicit intervention, therapist 2). Additionally, therapists described that participants tended to use explicit rules that they may have acquired through care givers and through previous therapy sessions. For example, they mentioned that participants had a tendency to develop explicit rules for different reasons, such as “*I think due to the classic (explicit) way of learning/improving gait*” (implicit intervention, therapist 3), ‘*especially due to instructions that have been used in earlier phases of rehabilitation*’ (explicit intervention, therapist 1) and ‘*A lot was made explicit by the former therapists, partner or daughter*’ (implicit intervention, therapist 3). In response to future use of the learned instructions (question 4), therapists thought that some participants who received the implicit intervention would probably switch back to the use of explicit rules.

#### Observed changes

A variety of observed changes after the intervention were reported by the therapists ranging from improvements in “spatiotemporal parameters” (e.g., better foot clearance, faster walking, or a better step through gait) to changes in “cognitions and emotions” (e.g., more confidence in walking, ease of walking and more joy in walking).

### Patient experiences with regard to feasibility

Table 5 represents the answers on participant questions related to their experiences with the received intervention. Overall, the participants were satisfied about the intervention and found the received instructions relatively easy to use.

**Table 5.**
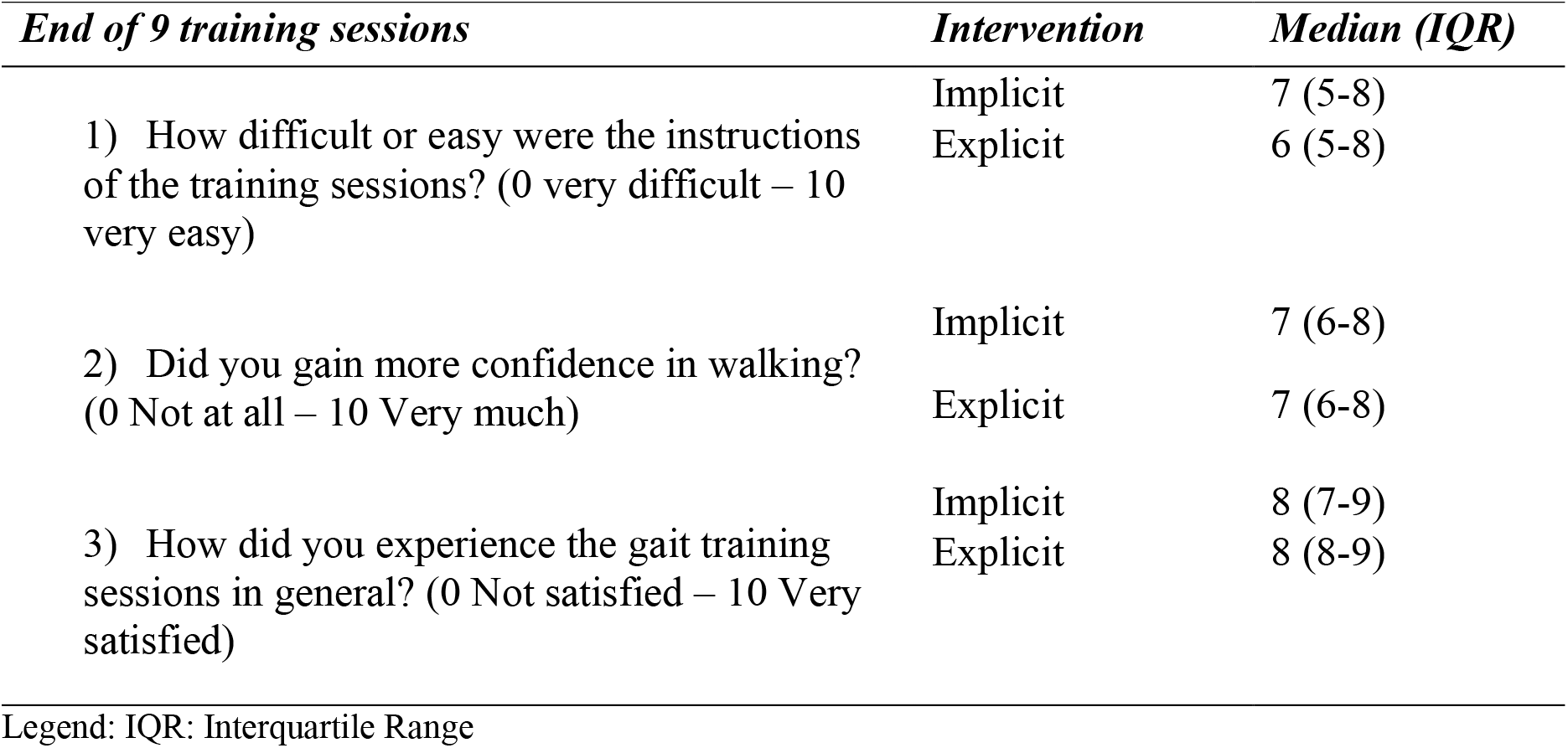
Responses of the patient questionnaire on the intervention’s feasibility

## Discussion

The aim of this process evaluation was to assess whether the implicit and explicit motor learning walking interventions for people after stroke delivered in a randomized controlled trial were performed as intended (fidelity) and to report the therapists and participants experiences with regard to feasibility. Overall, the interventions were delivered as intended with regard to fidelity and therapist and participants were generally positive about the feasibility of both interventions.

### Methodological quality, strength and limitations of the study

Within the process evaluation three factors were observed that may have influenced contrast between the implicit and explicit interventions. First, therapists were exposed to knowledge of both interventions, and needed to constantly switch between interventions, which may have led to contamination. Providing both interventions may also be seen as a strength of the study as in real life therapists equally need to switch between different learning approaches. Second, therapist observed that participants had the tendency to self-develop explicit rules. They mentioned that some participants tried to consciously control their movements, which is a common characteristic of people after stroke [18,19]. Furthermore, therapists reported that explicit rules may also have been acquired through support of caregivers and earlier received therapy sessions (prior to the study). The presence and possible use of explicit rules, may have caused the implicit intervention to lean more towards the middle of the implicit-explicit continuum. Third, therapists were more reserved in the provision of explicit rules in people with lower cognitive abilities which in turn this may also have contributed to a diminished the contrast between interventions. Despite good protocol adherence the process evaluation also showed that it is difficult to keep interventions 100% implicit or explicit. Incidentally (14% of the implicit sessions; 1% of the explicit sessions) implicit instructions were provided within the explicit intervention and vice versa. Also, in some cases the number of explicit rules within the explicit interventions was very low.

A strength of the study was that the interventions and population reflect real life practice very well as the interventions took place in the home environment of the participants and implicit and explicit instructions were tailored towards the individual walking problem. Also, the identified factors that possibly influenced the contrast of the interventions, should be taken into account and cannot be ignored when translating findings to daily life practice. The high intervention validity, and positive experiences by therapists and participants contribute to the generalizability and transferability of findings to the overall stroke population and clinical practice.

### Results compared to other studies

To our knowledge this was the first study to examine the fidelity and experiences of an implicit and explicit motor learning walking intervention that took place in the home environment of people after stroke. Nonetheless, some insights can be gained from studies in applied sports settings e.g., [12,20]. The average number of explicit rules, five in total, may in first instance seem remarkably low compared to studies that used eight [20] to twelve [12] explicit rules. However, in order to optimize the ecological validity and implications for clinical practice, we used personalized instructions rather than a fixed amount of and predetermined content for the instructions. As, extensively discussed by Bobrownicki et al [21], within future research the set of explicit instructions, with regard to quantity and quality should carefully be selected. More explicit rules may not necessarily be better and it may not always reflect real-life practice.

Therapists not only observed changes related to walking speed (main outcome parameter of the trial) but also reported changes in other spatiotemporal gait, feelings of joy and confidence. Similar findings were observed in another process evaluation that evaluated a movement imagery intervention in people after stroke [22]. Next to the primary outcome a variety of potential benefits e.g., related to confidence, were reported by the therapists. Future studies may consider incorporating different outcome measures that cover a broader range of improvements.

### Implications for research and clinical practice

From the process evaluation various implications for research and clinical practice can be learned. Firstly, therapists experienced that partners or family members, with good intensions used explicit rules (thus indirectly violated the protocol) to explain how an analogy should be performed. Therefore, it is important to have similar views and standardize treatments (i.e., mutual understanding of implicit learning) with caregivers and members of the multidisciplinary team who treat the patient.

Furthermore, finding a suitable (meaningful) analogy may require time and the process of developing a successful analogy may take longer than one therapy session. It can be a creative process as the analogy should reflect correct movement pattern and preferably be meaningful to the participant [8]. Examples of analogies to facilitate walking are reported (Table 3), also a few other analogies to facilitate gait have been described by Kleynen et al [8] and Jie et al [3]. We hope that sharing the delivered analogies (Table 3) may function as a source of inspiration

Tailored implicit and explicit motor learning walking interventions can be applied in a feasible manner to people after stroke who are in the chronic phase of recovery. However, various factors such as cognitive (in)abilities or the tendency to self-develop explicit rules can influence the implicit or explicit nature of the intervention.

### Conclusion

The delivery of the implicit and explicit motor learning walking interventions were successful in terms of fidelity. Within the process evaluation three factors were observed that may have influenced contrast between the implicit and explicit interventions e.g., less explicit rules were provided in people with lower cognitive abilities. Therapists and participants were generally positive about the intervention’s feasibility.

## Data Availability

The data that support the findings of this study are available from the corresponding author, upon a reasonable request.

## Clinical Trial Registration

The trial was registered at the [Netherlands Trial Registry](https://www.trialregister.nl/) (NL6133)

## Acknowledgements

This work was supported by Nationaal Regieorgaan Praktijkgericht Onderzoek SIA (RAAKPRO; grant number 2014-01-49PRO). We thank Marloes Jansen and Marius Wientgen for their support in data collection and data processing.

